# The Overlooked Burden: Prevalence, Risk Factors, and Neonatal Outcomes of Fetal Macrosomia from a Prospective Pregnancy Cohort in Sub-Saharan Africa

**DOI:** 10.64898/2026.11.26.26369058

**Authors:** Yahaya Idris, Milly Wilson, Hawanatu Jah, Marie-Laure Volvert, Baboucarr Njie, Rachel Craik, Fatoumata Kongira, Joseph Akuze, Anifa Valá, Moses Mukhanya, Angela Koech, Marleen Temmerman, Esperança Sevene, Peter von Dadelszen, Umberto D’Alessandro, Hannah Blencowe, Anna Roca, the PRECISE Network

**Author notes:** Joint senior authors (Hannah Blencowe, Anna Roca). **Corresponding author**: Yahaya Idris.

## Abstract

Macrosomia (birthweight ≥4000 g) is associated with adverse maternal and neonatal outcomes, with extreme macrosomia (≥4500 g) carrying the greatest risk. Most evidence on macrosomia comes from high-income countries, while data from sub-Saharan Africa, a region with unacceptably high preventable maternal and child morbidity and mortality, remain scarce.

We conducted a secondary analysis of data from the PREgnancy Care Integrating translational Science Everywhere prospective cohort in The Gambia, Kenya and Mozambique (2019-2022). We recruited pregnant women aged 16-49 years through antenatal clinics at seven government-run health facilities and collected data at scheduled visits. We used descriptive and logistic regression analyses to estimate prevalence, risk factors, and neonatal outcomes, and to compare outcomes across countries and between macrosomia strata (mild 4000g - <4500 g vs extreme ≥4500 g). Variables with p-values ≤ 0.1 in bivariate analyses were included in the multivariate model, with statistical significance defined as p-value < 0.05.

Among 5,814 births, 2.1% (n=122) were macrosomia overall and 0.2% (n=10) were extreme macrosomia. Gestational age (GA) >42 weeks (adjusted odds ratio (aOR): 2.65 [95% confidence interval (CI) 1.61-4.38]), parity 2-4 (aOR: 1.58 [1.02-2.43]) and ≥5 (aOR: 3.35 [1.78-6.32]), mid-upper arm circumference ≥30cm (aOR: 2.85 [1.72-4.71]) and maternal height ≥1.60m (aOR: 2.19 [1.47-3.26]) were associated with macrosomia, while female sex was protective (aOR: 0.60 [0.41-0.88]). Macrosomia increased the risk of C-section (OR: 2.62 [1.56-4.40]) and stillbirth (OR: 3.40 [1.15-10.08]). Extreme macrosomia was associated with a 4-fold and 13-fold increased risk of Caesarean birth (OR: 4.05 [1.06-15.50]) and postpartum admission (OR: 13.29 [1.58-111.89]), respectively. In the entire cohort, macrosomia accounted for 4.8% of deliveries by Caesarean birth, 3.6% of birth asphyxia, 3.3% of post-delivery admission, 3.5% of stillbirths, and 7.1% of neonatal deaths.

In sub-Saharan Africa, macrosomia is associated with a substantial risk of stillbirth. Identified modifiable risk factors for macrosomia, including obesity and post-term pregnancy, could be addressed through health education, lifestyle modification and labour induction (if accurate GA estimation is guaranteed). Intrapartum stillbirth, neonatal deaths and morbidities could be averted through improved intrapartum management.

## INTRODUCTION

Globally, approximately 9% of neonates are born with macrosomia (≥4000 g). Reported prevalence varies widely, ranging from 5-20% in high-income countries (HICs) to 0.5-14.9% in low-and middle-income countries (LMICs), including those in sub-Saharan Africa (SSA)[1,2]. For extreme macrosomia (≥4500 g), a prevalence of 1.2% has been reported [3].

Macrosomia is associated with increased short- and long-term health risks for both mothers and infants[4]. For mothers, delivering a macrosomic baby increases the risk of labour complications, operative delivery, postpartum haemorrhage, and postpartum morbidity, with potential long-term pelvic floor and anal sphincter injury [1,2,5–7]. In infants, short-term complications include perinatal mortality, birth trauma, and metabolic disturbances, while long-term health consequences include childhood obesity and metabolic disorders, such as diabetes [1,2,5,6,8].

Several maternal risk factors increase the risk of macrosomia. Modifiable risk factors include excessive gestational weight gain, obesity, diabetes, and post-term delivery [2,9,10]. Non-modifiable factors include advanced maternal age, male fetal sex, multiparity and maternal height [1,6,11].

Currently, Africa is undergoing a socio-demographic, epidemiological, nutritional, and economic transition, alongside rapid urbanisation. These shifts are accompanied by rising rates of obesity and diabetes [12–14], which are expected to increase the prevalence of macrosomia and related adverse outcomes [1,15]. Despite global research on macrosomia, data from SSA, a region with unacceptably high preventable maternal and child morbidity and mortality, remain limited. To date, only one multi-country study based on hospital records with a cross-sectional design and conducted more than a decade ago has been identified [1].

To address this data gap, we present findings from a prospective multi-country pregnancy cohort study from the PREgnancy Care Integrating translational Science, Everywhere (PRECISE) cohort, which includes more than 6,000 pregnant women in SSA (The Gambia, Kenya and Mozambique). Our study evaluates the prevalence, risk factors and perinatal or neonatal outcomes of macrosomia.

## MATERIALS AND METHODS

### Ethical Consideration

The PRECISE study received ethical approval from the Research Ethics Committee at King’s College London (Ref HR-17/18–7855) as well as from local ethics committees of the respective partner institutions in The Gambia (Ref SCC 1619), Kenya (Ref 2018/REC-74), and Mozambique (545/CNBS/18). Participants enrolled in the study were provided with information in a language and format they could understand to support informed decision-making. Written informed consent was obtained through either a signature or a thumbprint before participation.

### Study Design and Settings

This study is a secondary analysis of data from the PRECISE pregnancy study, a prospective cohort of pregnant women recruited between 2019 and 2022 across The Gambia (Farafenni district), Kenya (Mariakani and Rabai sub-counties), and Mozambique (Manhiça and Xinavane districts). The study investigates placental disorders, including pregnancy hypertension, fetal growth restriction (FGR), preterm birth, and stillbirth. Details of the study protocol and cohort have been published elsewhere [16,17].

### Study Population and Data Collection

The study participants were pregnant women aged 16-49 years who provided consent and were recruited into the PRECISE pregnancy cohort. We enrolled them through antenatal care (ANC) clinics at participating facilities in each country. Participants were followed across up to four scheduled visits, including: (i) enrolment (<28 weeks’ gestation), (ii) second ANC visit (≥28 weeks’ gestation and at least 8 weeks after enrolment visit), (iii) delivery (onset of labour), and (iv) a last visit for mother and infant within six weeks postpartum. At each visit, we collected socio-demographic, obstetric, and other relevant clinical data along with biological samples.

Baseline maternal data used in this analysis, such as socio-demographic and nutritional characteristics, height, MUAC, parity, history of C-section, and GA, were collected at the enrolment visit using interviewer-administered questionnaires. Birth outcome data, such as birth weight (measured with the newborn unclothed), were measured immediately after delivery in labour wards of participating health facilities in each country. Study midwives and nurses collected most data and entered it immediately into an electronic database, keeping a paper record for data quality control and future reference. Hospital facility nurses and midwives assessed Apgar scores, and these were subsequently entered into the electronic database by the study staff. We determined GA at enrolment and at the end of pregnancy using a combination of methods and algorithms. Briefly, these included, in order of priority, use of routine ultrasound scan (accounted for 40%), measurements by the trained study staff (including clinicians in The Gambia and Kenya) using a digital device “TraCer” (19.3%), and last menstrual period (28.1%) and clinical symphysis-fundal height (10.7%) when the former two were unavailable. (Further details are presented in Supporting Information (S) **S1 Table**)[17]. Postpartum data included in this analysis were collected within 48 hours of delivery and up to 28 days during the six-week postpartum visit.

### Study Materials and Data Quality Control

Data were collected electronically with tablets equipped with ODK-X or REDCap Apps. Weight and height (length) measurements for both mothers and neonates were taken using Seca scales and a Shakir tape for mid-upper arm circumference (MUAC). These anthropometric measurements were recorded at least twice to minimise the observer’s error. A margin of error of <0.5 was acceptable between two measurements. Where this margin was >0.5, repeat measurement(s) were done until the desired precision of <0.5 was obtained and then recorded. Anthropometric standardisations were conducted every 3-6 months, and staff were retrained as necessary across study sites. An innovative TraCer device, connected to an app installed on Android devices and requiring minimally trained personnel, was used by the study staff in The Gambia and Kenya only to estimate GA, as routine ultrasound scans were widely available in Mozambique (82.3% availability) [18]. Data collected in each country were synchronised and stored daily in the central database. Data were checked for completeness, consistency, accuracy, and precision, and cleaning was performed as an ongoing process across all sites supported by a central data team.

### Outcome Definitions and Classification Criteria

Macrosomia was defined as a birthweight of ≥4000 g. Macrosomia was further divided into mild macrosomia (birth weight between ≥4000 g and <4500 g) and extreme (≥4500 g). The comparison group for macrosomia was defined as normal birth weight (2500 g to <4000 g).

Stillbirth was defined as fetal demise occurring at ≥20^+0^ weeks of gestation before birth, while neonatal death refers to the death of a liveborn baby within 28 days of birth. For this study, neonates with a 5-minute Apgar score of less than 7 were considered to have intrapartum-related asphyxia, subclassified as moderate (4 to 6) or severe (less than 4). Neonatal intensive care unit (NICU) or post-delivery hospital admission was considered when hospitalisation occurred following delivery, separate from the mother.

### Participant Inclusion and Exclusion

The eligibility criteria set out by the parent study were pregnant women aged 16-49 years, attending ANC in a designated collaborating health facility and not a referral from another facility. For this sub-study, our eligibility criteria (**Fig 1**) are: Inclusion: (1) Prevalence analysis: Recorded pregnancy outcome (stillbirth, live birth, singleton or multiple gestation, low birthweight (LBW; < 2500 g), normal birthweights and macrosomia) (2) Bivariate/multivariate analysis: All recorded singleton gestations, normal birthweights and macrosomia. Exclusion: (1) All analyses: Miscarriages <20^+0^ weeks of gestation, missing birth records; (2) Bivariate/multivariate analysis: Multiple gestations and LBW because they constituted another risk group for poor perinatal outcomes and, therefore, might have confounded our findings.

**Fig 1:**
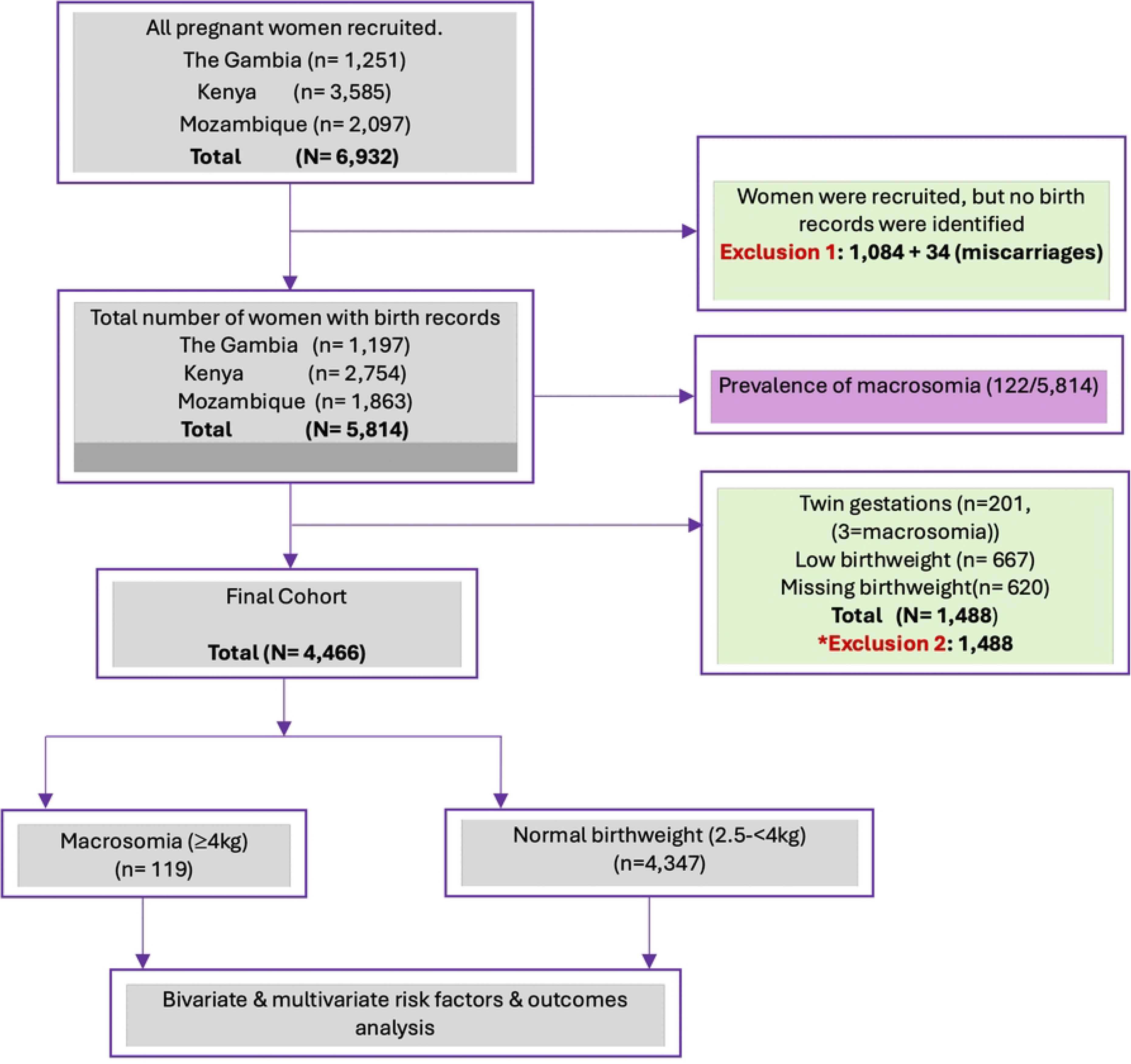
Fetal Macrosomia Study Profile from the PRECISE (PREgnancy Care Integrating translational Science Everywhere) Pregnancy Cohort in The Gambia, Kenya and Mozambique. *Exclusions are not mutually exclusive from each other; for example, twin gestations could also be in the categories with missing data.

### Statistical Analysis

Baseline characteristics of the cohort were described using median [IQR] for quantitative variables and frequencies, with percentages for categorical variables. The prevalence of macrosomia was estimated for each country and overall, and expressed as percentages. Prevalence analysis was computed as the number of macrosomic newborns divided by total births, irrespective of outcomes.

To explore factors associated with macrosomia, we developed an original conceptual framework based on a literature review [1,2,8,14,19] and the consideration of clinically and biologically plausible pathways of macrosomia in low-resource settings (**Fig 2**). Variables in the conceptual framework available in our study are highlighted in yellow.

**Fig 2:**
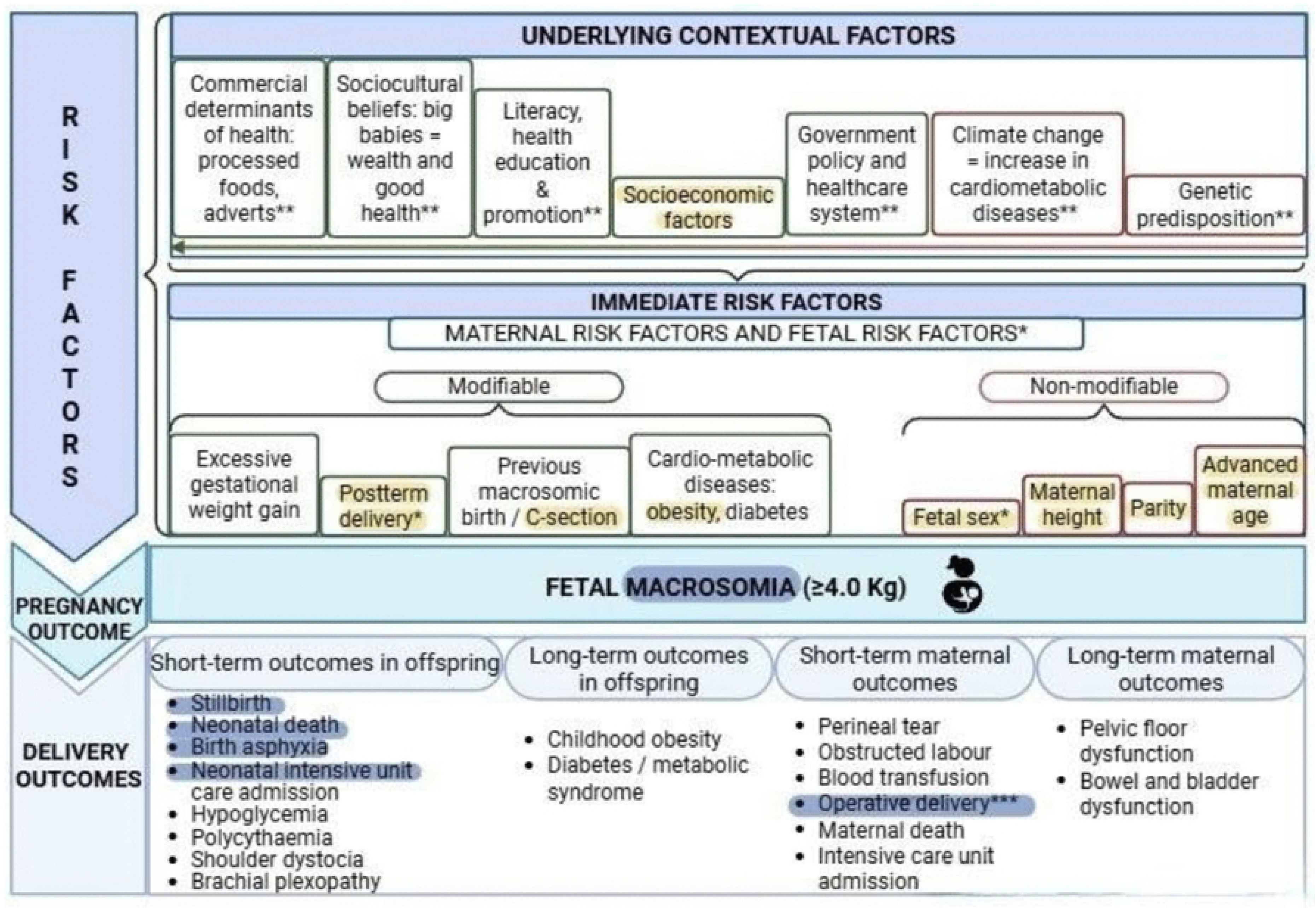
A Conceptual Framework of the Risk Factors for Fetal Macrosomia and Associated Maternal and Perinatal Short- and Long-term Outcomes. * Fetal risk factors. ** Not commonly reported in the literature or emerging. *** Strictly a maternal outcome but included as part of neonatal outcome analysis. Coded yellow & blue: Available variables included in the risk factors & outcomes analysis, respectively. Coded red: non-modifiable risk factors. Coded green: Modifiable risk factors.

The conceptual framework guided our choice of variables for the univariate analysis and informed the adjusted analysis. Variables with a p-value ≤0.1 in the univariate analysis were included in the multivariate model. Country of residence was forced into the multivariate model because we perceived it as a strong potential confounder. In all univariate analyses, including the outcome analysis below, effect sizes were reported as odds ratios (ORs) and as adjusted odds ratios (aORs) in the multivariate model at 95% confidence intervals (CIs). A p-value of <0.05 was considered statistically significant in the multivariate risk factors and outcome analyses.

A univariate analysis was conducted to assess the perinatal or neonatal outcomes (highlighted in blue in the conceptual framework as the available covariates, **Fig 2**) as outcome variables against the dichotomised exposure variable (macrosomia vs normal birthweight). Stratified analyses for the severity of macrosomia (birthweights of < 4500 g vs ≥4500 g) and country were conducted using logistic regression models for the same set of variables as in the crude analysis.

Missing data were addressed by multiple imputation, using the Multivariate Imputation by Chained Equations (MICE) package in R statistical software (v4.1.2; R Core Team 2021). Twenty imputed datasets were generated, using all prognostic variables and outcomes. Results were pooled using Rubin’s rules[20]. All analyses were conducted using R statistical software version 4·1·2.

## RESULTS

### Baseline characteristics

We included a total of 5,814 births in our analysis (**Fig 1**). The median maternal age at enrolment was 25 years, and the median birthweight was 3100 g. Additional descriptive statistics are provided in **S2 and S3 Tables**. The overall prevalence of macrosomia in the cohort was 2.1% (122 of 5,814), ranging from 1.7% in The Gambia to 2.1% in Kenya and 2.4% in Mozambique. The overall prevalence of mild macrosomia was 1.9% (112 of 5,814), and extreme macrosomia was 0.2% (10 of 5,814).

### Risk factors analysis

A total of 4,466 births (76.8%) out of 5814 birth records were included in the risk factors analysis, 119 with macrosomia (2.7%). In the crude analysis, GA at delivery, parity, maternal MUAC, height and age, and country were all positively associated with macrosomia, while female sex was protective (**Table 1**). In the multivariate analysis, all these variables, except maternal age and country, remained independently associated with macrosomia (aOR = 2.65 [95%CI 1.61-4.38] for GA > 42 weeks, aOR = 1.58 [95%CI 1.02-2.43] and aOR = 3.35 [95%CI 1.78-6.32] for parities of 2-4 and ≥5 (vs 0-1), aOR = 2.85 [95%CI 1.72-4.71] for MUAC of ≥30cm (vs <30cm), and aOR = 2.19 [95%CI 1.47-3.26] for maternal height of ≥1.60m (vs <1.60m)). Female sex remained protective (aOR = 0.60 [95%CI 0.41-0.88]).

**Table 1:**
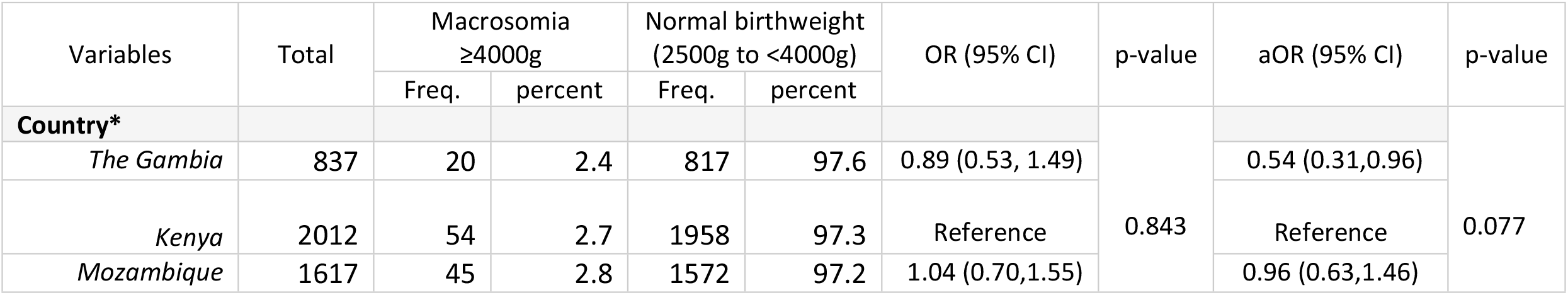
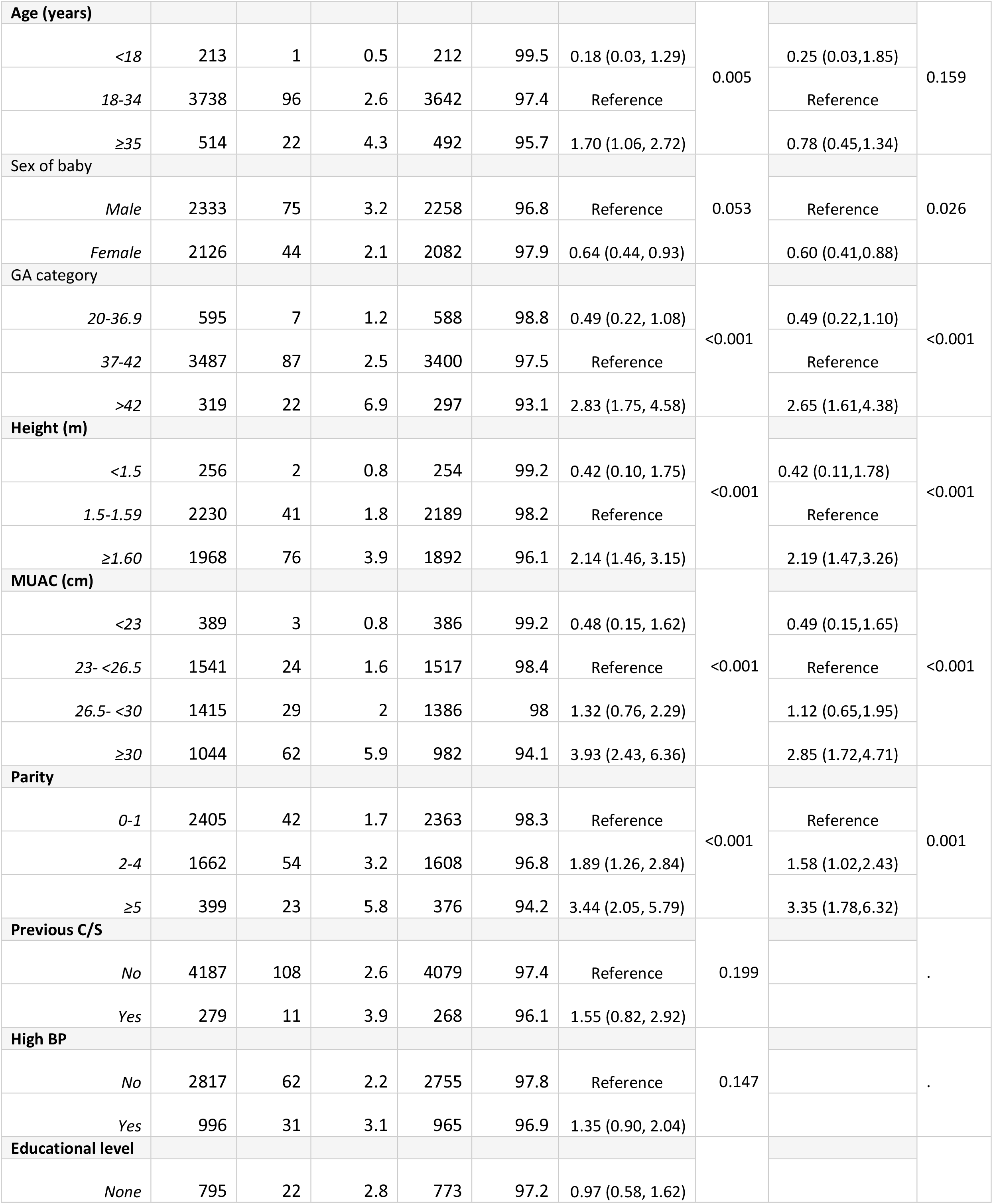
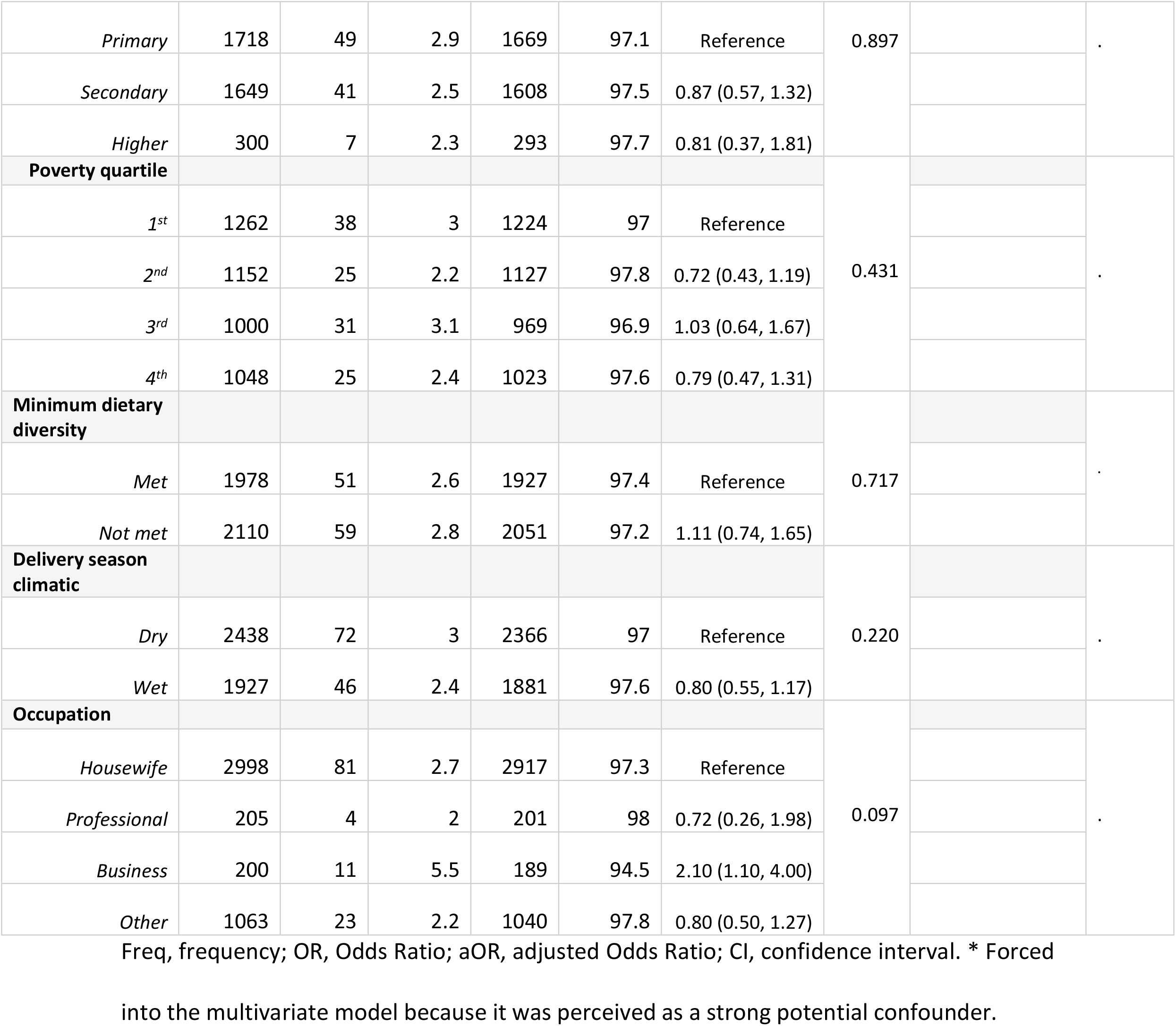
Bivariate and Multivariate Regression Analysis of Risk Factors for Fetal Macrosomia from PRECISE (PREgnancy Care Integrating translational Science Everywhere) Pregnancy Cohort in The Gambia, Kenya and Mozambique.

| Variables | Total | Macrosomia ≥4000g |  | Normal birthweight (2500g to <4000g) |  | OR (95% CI) | p-value | aOR (95% CI) | p-value |
|---|---|---|---|---|---|---|---|---|---|
|  |  | Freq. | percent | Freq. | percent |  |  |  |  |
| **Country\*** |  |  |  |  |  |  | 0.843 |  | 0.077 |
| *The Gambia* | 837 | 20 | 2.4 | 817 | 97.6 | 0.89 (0.53, 1.49) |  | 0.54 (0.31,0.96) |  |
| *Kenya* | 2012 | 54 | 2.7 | 1958 | 97.3 | Reference |  | Reference |  |
| *Mozambique* | 1617 | 45 | 2.8 | 1572 | 97.2 | 1.04 (0.70,1.55) |  | 0.96 (0.63,1.46) |  |
| **Age (years)** |  |  |  |  |  |  | 0.005 |  | 0.159 |
| *<18* | 213 | 1 | 0.5 | 212 | 99.5 | 0.18 (0.03, 1.29) |  | 0.25 (0.03,1.85) |  |
| *18-34* | 3738 | 96 | 2.6 | 3642 | 97.4 | Reference |  | Reference |  |
| *≥35* | 514 | 22 | 4.3 | 492 | 95.7 | 1.70 (1.06, 2.72) |  | 0.78 (0.45,1.34) |  |
| Sex of baby |  |  |  |  |  |  | 0.053 |  | 0.026 |
| *Male* | 2333 | 75 | 3.2 | 2258 | 96.8 | Reference |  | Reference |  |
| *Female* | 2126 | 44 | 2.1 | 2082 | 97.9 | 0.64 (0.44, 0.93) |  | 0.60 (0.41,0.88) |  |
| GA category |  |  |  |  |  |  | <0.001 |  | <0.001 |
| *20-36.9* | 595 | 7 | 1.2 | 588 | 98.8 | 0.49 (0.22, 1.08) |  | 0.49 (0.22,1.10) |  |
| *37-42* | 3487 | 87 | 2.5 | 3400 | 97.5 | Reference |  | Reference |  |
| *>42* | 319 | 22 | 6.9 | 297 | 93.1 | 2.83 (1.75, 4.58) |  | 2.65 (1.61,4.38) |  |
| **Height (m)** |  |  |  |  |  |  | <0.001 |  | <0.001 |
| *<1.5* | 256 | 2 | 0.8 | 254 | 99.2 | 0.42 (0.10, 1.75) |  | 0.42 (0.11,1.78) |  |
| *1.5-1.59* | 2230 | 41 | 1.8 | 2189 | 98.2 | Reference |  | Reference |  |
| *≥1.60* | 1968 | 76 | 3.9 | 1892 | 96.1 | 2.14 (1.46, 3.15) |  | 2.19 (1.47,3.26) |  |
| **MUAC (cm)** |  |  |  |  |  |  | <0.001 |  | <0.001 |
| *<23* | 389 | 3 | 0.8 | 386 | 99.2 | 0.48 (0.15, 1.62) |  | 0.49 (0.15,1.65) |  |
| *23- <26.5* | 1541 | 24 | 1.6 | 1517 | 98.4 | Reference |  | Reference |  |
| *26.5- <30* | 1415 | 29 | 2 | 1386 | 98 | 1.32 (0.76, 2.29) |  | 1.12 (0.65,1.95) |  |
| *≥30* | 1044 | 62 | 5.9 | 982 | 94.1 | 3.93 (2.43, 6.36) |  | 2.85 (1.72,4.71) |  |
| **Parity** |  |  |  |  |  |  | <0.001 |  | 0.001 |
| *0-1* | 2405 | 42 | 1.7 | 2363 | 98.3 | Reference |  | Reference |  |
| *2-4* | 1662 | 54 | 3.2 | 1608 | 96.8 | 1.89 (1.26, 2.84) |  | 1.58 (1.02,2.43) |  |
| *≥5* | 399 | 23 | 5.8 | 376 | 94.2 | 3.44 (2.05, 5.79) |  | 3.35 (1.78,6.32) |  |
| **Previous C/S** |  |  |  |  |  |  | 0.199 |  | . |
| *No* | 4187 | 108 | 2.6 | 4079 | 97.4 | Reference |  |  |  |
| *Yes* | 279 | 11 | 3.9 | 268 | 96.1 | 1.55 (0.82, 2.92) |  |  |  |
| **High BP** |  |  |  |  |  |  | 0.147 |  | . |
| *No* | 2817 | 62 | 2.2 | 2755 | 97.8 | Reference |  |  |  |
| *Yes* | 996 | 31 | 3.1 | 965 | 96.9 | 1.35 (0.90, 2.04) |  |  |  |
| **Educational level** |  |  |  |  |  |  |  |  |  |
| *None* | 795 | 22 | 2.8 | 773 | 97.2 | 0.97 (0.58, 1.62) |  |  |  |
| *Primary* | 1718 | 49 | 2.9 | 1669 | 97.1 | Reference | 0.897 |  | . |
| *Secondary* | 1649 | 41 | 2.5 | 1608 | 97.5 | 0.87 (0.57, 1.32) |  |  |  |
| *Higher* | 300 | 7 | 2.3 | 293 | 97.7 | 0.81 (0.37, 1.81) |  |  |  |
| **Poverty quartile** |  |  |  |  |  |  | 0.431 |  | . |
| *1<sup>st</sup>* | 1262 | 38 | 3 | 1224 | 97 | Reference |  |  |  |
| *2<sup>nd</sup>* | 1152 | 25 | 2.2 | 1127 | 97.8 | 0.72 (0.43, 1.19) |  |  |  |
| *3<sup>rd</sup>* | 1000 | 31 | 3.1 | 969 | 96.9 | 1.03 (0.64, 1.67) |  |  |  |
| *4<sup>th</sup>* | 1048 | 25 | 2.4 | 1023 | 97.6 | 0.79 (0.47, 1.31) |  |  |  |
| **Minimum dietary diversity** |  |  |  |  |  |  | 0.717 |  | . |
| *Met* | 1978 | 51 | 2.6 | 1927 | 97.4 | Reference |  |  |  |
| *Not met* | 2110 | 59 | 2.8 | 2051 | 97.2 | 1.11 (0.74, 1.65) |  |  |  |
| **Delivery season climatic** |  |  |  |  |  |  | 0.220 |  | . |
| *Dry* | 2438 | 72 | 3 | 2366 | 97 | Reference |  |  |  |
| *Wet* | 1927 | 46 | 2.4 | 1881 | 97.6 | 0.80 (0.55, 1.17) |  |  |  |
| **Occupation** |  |  |  |  |  |  | 0.097 |  | . |
| *Housewife* | 2998 | 81 | 2.7 | 2917 | 97.3 | Reference |  |  |  |
| *Professional* | 205 | 4 | 2 | 201 | 98 | 0.72 (0.26, 1.98) |  |  |  |
| *Business* | 200 | 11 | 5.5 | 189 | 94.5 | 2.10 (1.10, 4.00) |  |  |  |
| *Other* | 1063 | 23 | 2.2 | 1040 | 97.8 | 0.80 (0.50, 1.27) |  |  |  |

Freq, frequency; OR, Odds Ratio; aOR, adjusted Odds Ratio; CI, confidence interval. \* Forced into the multivariate model because it was perceived as a strong potential confounder.

### Perinatal and neonatal outcomes among children with macrosomia

Neonates with macrosomia (vs normal weight newborns) were at increased risk of Caesarean births (OR = 2.62 [95%CI 1.56-4.40]) and stillbirth (OR = 3.40 [95%CI 1.15-10.08]) (**Table 2**).

**Table 2:** Univariate Analysis of Fetal Outcomes of Macrosomia from the PRECISE (PREgnancy Care Integrating translational Science Everywhere) Pregnancy Cohort in The Gambia, Kenya and Mozambique.

| Variables | Total<br>n= (%) | Macrosomia<br>≥4000g | Normal birthweight<br>(2500g to <4000g) | OR (95% CI) | p-value |
|---|---|---|---|---|---|
|  |  | n (%) | n (%) |  |  |
| Mode of delivery | N=4413 | n=116 | n=4297 | - | - |
| *Vaginal (assisted, unassisted, breech)* | 3951 (89.5) | 90 (77.6) | 3861 (89.9) | Reference | - |
| *Caesarean section* | 462 (10.5) | 26 (22.4) | 436 (10.1) | 2.62 (1.56, 4.40) | <0.001 |
| Birth Asphyxia (5-min) | N=4131 | n=104 | n=4027 |  |  |
| *≥7* | 4045 (97.9) | 100 (96.2) | 3945 (98.0) | Reference | - |
| *<7* | 86 (2.1) | 4 (3.8) | 82 (2.0) | 1.94 (0.67,5.62) | 0.223 |
| Post-delivery admission | N=4188 | 104 | 4084 |  |  |
| *No* | 4122 (98.4) | 100 (96.2) | 4022 (97.6) | Reference | - |
| *Yes* | 66 (1.6) | 4 (3.8) | 62 (1.5) | 2.11 (0.72,6.18) | 0.172 |
| Stillbirth | N=4466 | n=119 | n=4347 |  |  |
| No | 4416 (98.9) | 115 (96.6) | 4301 (98.9) | Reference | - |
| Yes | 50 (1.1) | 4 (3.4) | 46 (1.1) | 3.40 (1.15,10.08) | 0.027 |
| Neonatal death | N=4466 | n=119 | n=4347 |  |  |
| *No* | 4450 (99.6) | 117 (98.3) | 4333 (99.7) | Reference | - |
| *Yes* | 16 (0.4) | 2 (1.7) | 14 (0.3) | 4.25 (0.83, 21.76) | 0.082 |

OR, Odds Ratio; CI, confidence interval.

Macrosomia accounted for 12.5% (2 out of 16) of neonatal deaths, 8.0% (4 out of 50) of stillbirths, 6.1% (4 out of 66) of post-delivery admissions, 5.6% (26 out of 462) of Caesarean births, and 4.7% (4 out of 86) of birth asphyxia cases recorded in the cohort. When we consider the entire cohort, including the LBW neonates, macrosomia accounted for 7.1% of neonatal deaths (2/28), 4.8% (26/547) of Caesarean births, 3.6% of birth asphyxia cases (4/111), 3.5% of stillbirths (4/115), and 3.3% of post-delivery admissions (4/120). Further details are provided in **Table 3**.

**Table 3:**
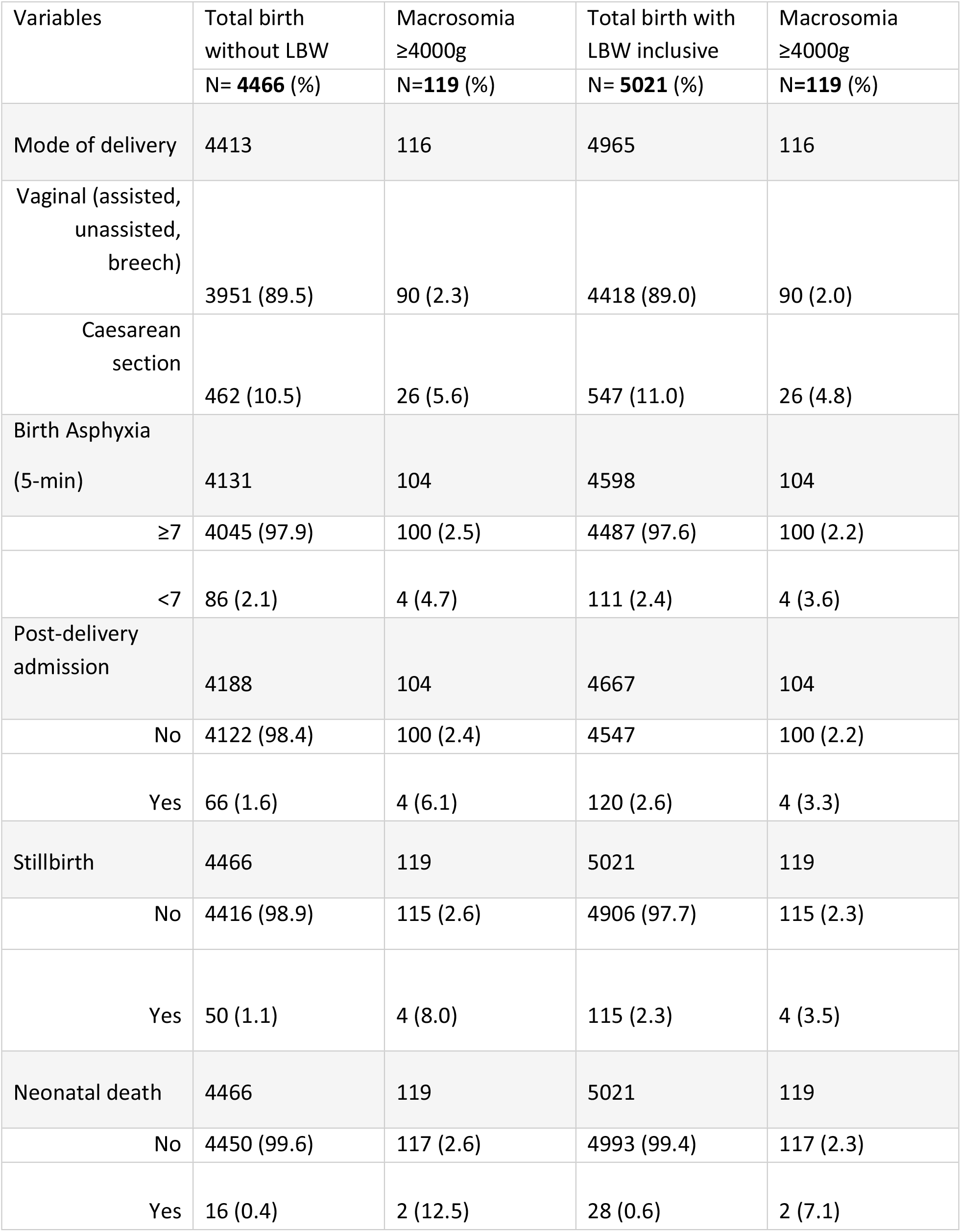
Percentage Contribution of Macrosomia to Perinatal/Neonatal Burdens from the PRECISE (PREgnancy Care Integrating translational Science Everywhere) Pregnancy Cohort in The Gambia, Kenya and Mozambique.

We conducted a subgroup analysis stratified by country; there was no strong evidence of statistical difference in the above-studied outcomes among the three countries (**S5 Table**).

Stratifying by severity (birthweights of 4000 - <4500 g vs ≥4500 g), neonates with extreme macrosomia (≥4500 g) had an increased risk of Caesarean birth (OR: 4.05 [95% CI 1.06 -15.50]) and post-delivery admission (OR: 13.29 [95% CI 1.58 -111.89]) compared with those with mild macrosomia (4000 - <4500 g) (**S4 Table**).

## Discussion

In this large multi-country study conducted in SSA, represented by The Gambia, Kenya and Mozambique, macrosomia affected approximately one in 50 newborns, with expected slight variations across countries, ranging from 1.7% in The Gambia to 2.4% in Mozambique, while severe macrosomia occurred in less than 0.5% of births. Macrosomic newborns had a substantially increased risk of adverse perinatal outcomes, particularly Caesarean birth and stillbirth. We identified several maternal factors associated with macrosomia that are potentially modifiable or can be identified during pregnancy, including GA, higher MUAC, parity and high maternal height.

The prevalence estimates in our cohort fall within the lower range of 2.0-14.9% reported in previous African studies [1,21,22]. Higher prevalences reported in these mostly hospital-based tertiary institutions in urban populations in Algeria (14.9%), Uganda (8.7%), Nigeria (7.5%), Egypt (4.6%), Morocco (4.4%) and Kenya (3.6%) likely reflect referrals of high-risk pregnancies combined with a greater burden of maternal obesity and diabetes (known risk factors for macrosomia) [13,14]. Our cohort was predominantly drawn from rural and peri-urban populations, where these factors may be less common. The prevalence of extreme macrosomia in our cohort was slightly lower than the 1.2% (IQR of 0.7-2%) reported in a large population-based multi-country study [3]. This slight difference might be explained by the inclusion of predominantly middle- and high-income countries, where obesity and diabetes might be more prevalent[23], with no representation from LMICs, including SSA, likely due to data gaps.

Even though macrosomia constituted a small proportion of the overall population, macrosomic babies carried a disproportionate burden of adverse outcomes, representing between 5% and 13% of stillbirths, Caesarean births, birth asphyxia cases, neonatal deaths, and post-delivery admissions. Compared with normal birthweight infants, the 2-4-fold increased odds of adverse outcomes observed in macrosomic babies in our study are comparable to previous studies [6,24,25]. However, the overall prevalence of adverse outcomes was disproportionately high in our study. A prevalence of 4.2% of perinatal morbidity and mortality (stillbirth, birth asphyxia, NICU admission or referral, and neonatal death) among macrosomic neonates vs 3.3% in normal birthweight neonates in Africa was reported by a multi-continental study [1]. The higher burden observed in our study might be attributed to differences in study settings, as our study cohort was largely drawn from primary and secondary health care facilities in rural and peri-urban settings in SSA, where access to specialist obstetrics and perinatal services is minimal compared to this other study that mostly used tertiary-hospital-based data. The fact that the study was published more than a decade ago might also have implications. Currently, Africa is undergoing nutritional and socio-demographic transitions, which may contribute to increased prevalence of obesity and diabetes[13,26,27], major determinants of macrosomia, therefore leading to a consequential increase in perinatal/neonatal burdens. Although the increased risk of adverse outcomes of macrosomic babies is observed worldwide, the risk of mortality in macrosomic babies is disproportionately higher in SSA. A multi-continental study including middle- and high-income countries that excludes Africa reported a mortality risk ranging from 0.05 to 0.1% (0.5-1/1000 live births) among macrosomic neonates [3], compared with up to 13% in our study.

Understandably, the overall Caesarean birth rates in our study (10.9% in the entire cohort) fall within the WHO thresholds of 10-15% for all pregnancies [28]; only 22% of our macrosomic babies were born by Caesarean, substantially lower than those reported by other studies within Africa (26.9–61.1%) and >50% in Latin America and Asia[1,6,21,29,30]. These disparities may suggest under-identification of mothers who require operative deliveries or those who were allowed to undergo trial of labour without adequate Caesarean birth intervention when needed, likely contributing to the elevated rates of stillbirths and neonatal deaths among macrosomic babies. As expected, in newborns with extreme macrosomia, the risks of Caesarean birth and post-delivery admissions were markedly higher, with up to 50% and 20% of these babies, respectively, experiencing these outcomes. Previous studies have reported similar dose-response trends [25,31,32].

Strengthening health systems capacity through targeted training of healthcare workers to identify macrosomic pregnancies, alongside improved access to specialist obstetric and neonatal care in rural and peri-urban settings, is essential if advances towards Universal Health Coverage and SDG targets for reducing neonatal mortality are to be achieved equitably across all regions and settings. While these attributable differences might be immediately obvious, future studies should focus on exploring the relationships between the impact of training of the local health care workforce, reserving suspected macrosomic deliveries for experienced and skilled midwives, access to specialist obstetric and perinatologist services and macrosomia burdens. This will ensure evidence-based allocation of these human resources and equitable distribution across all tiers of health care service delivery, especially in resource-limited settings such as SSA.

Maternal obesity assessed using MUAC (previous studies mostly used BMI) and post-term pregnancy were the key modifiable factors associated with macrosomia in our cohort, consistent with previous evidence [1,33]. MUAC offers a practical and reliable alternative to BMI for assessing maternal nutritional status during pregnancy[34,35], particularly in LMICs, where pre-pregnancy weight is often unavailable. These findings underscore the need to systematically integrate targeted dietary counselling and promotion of physical activity into ANC programs [36]. Accurate assessment of GA is also critical, as a post-term pregnancy was associated with macrosomia in our study. Failure to identify prolonged gestation represents a missed opportunity for intervention. Timely recognition should prompt appropriate obstetric management, including timely induction of labour when indicated, as lower neonatal mortality risk has been observed among large for gestational age babies in countries with active induction protocols for post-term delivery[3]. Non-modifiable factors such as maternal height, fetal sex or multiparity were associated with macrosomia in our study, in line with previous findings [1,6]. Although non-modifiable, they provide important information for guiding risk stratification during ANC visits and inform intensified monitoring and preparedness for potential delivery complications.

This study has several important strengths. Its prospective design, large multi-country cohort and inclusion of rural and peri-urban populations enhance the generalisability of the findings. The use of a novel conceptual framework to guide the risk factors analysis strengthens the analytical approach and supports a holistic interpretation of the results. Stratification by macrosomia severity allowed exploration of gradients in adverse outcomes across birthweight categories, providing clinically relevant information. On the other hand, some limitations should also be acknowledged. The relatively small number of extremely macrosomic births limited statistical power for the stratified analysis. Missing birthweight data, largely attributable to COVID-19–related disruptions, may have introduced bias, as this would have reflected constraints of data collection under pandemic conditions. Finally, relevant outcomes such as neonatal hypoglycaemia, birth traumas, and risk factors like diabetes, a major determinant for macrosomia, were not assessed in the study cohort and could not be accounted for in the analysis.

## Conclusion

Macrosomia, although relatively uncommon in SSA, remains an important and mostly preventable contributor to adverse perinatal outcomes. Our findings call for explicit prioritisation within maternal and newborn health policies in SSA. ANC should be strengthened to identify high-risk pregnancies through improved GA assessment and routine screening for maternal obesity using feasible measures such as MUAC. Health systems should ensure timely access to obstetric care, including the capacity to manage suspected macrosomia or prolonged gestation, warranting an explicit call for future research to focus in this direction. Integrating these actions into existing care platforms would represent a scalable opportunity to reduce preventable adverse outcomes associated with macrosomic births in resource-limited settings.

## Data Availability

Data for scientific use could be accessed on formal request to the PREgnancy Care Integrating translational Science Everywhere (PRECISE) data access committee through the corresponding authors. Further information on data availability and access could be found at https://precisenetwork.org/.

## Acknowledgements

We are grateful to all the study participants and stakeholders, including the respective countries’ hospital staff, traditional birth attendants (The Gambia), community leaders, and government officials. We thank the field workers, nurses, laboratory staff, data management team, MRCG operations (research support office, transport team, finance, and other departments), Maria Climent and all members of the PRECISE Network.

## PRECISE Network Members

| **In-country teams** | **Members** |
|---|---|
| THE GAMBIA: Medical Research Council Unit The Gambia at the London School of Hygiene and Tropical Medicine, Fajara | Umberto D’Alessandro, Anna Roca, Hawanatu Jah, Andrew Prentice, Melisa Martinez-Alvarez, Brahima Diallo, Abdul Sesay, Sambou Suso, Baboucarr Njie, Fatima Touray, Yahaya Idris, Fatoumata Kongira, Modou F.S. Ndure, Gibril Gabbidon, Lawrence Gibba, Abdoulie Bah and Yorro Bah. |
| KENYA: Aga Khan University, Nairobi | Marleen Temmerman, Angela Koech, Patricia Okiro, Geoffrey Omuse, Grace Mwashigadi, Consolata Juma, Joseph Mutunga, Moses Mukhanya, Onesmus Wanje, Isaac Mwaniki, Marvin Ochieng, Emily Mwadime, Amina Abubakar |
| MOZAMBIQUE : Centro de Investigação em Saúde de Manhiça, Manhiça and Eduardo Mondlane University, Faculty of Medicine (FACMED) | CISM - Corssino Tchavana, Helena Boene, Lazaro Quimice, Sonia Maculuve, Inacio Mandomando, Abel Nhama, Chenjerai Jairoce<br>UEM/FACMED - Esperança Sevene, Anifa Valá, Carla Carrilho |
| **Central co-ordinating team** |  |
| Department of Women and Children’s Health, School of Life Course Sciences, Faculty of Life Sciences and Medicine, King’s College London | Peter von Dadelszen, Laura A. Magee, Rachel Craik, Marie-Laure Volvert, Hiten Mistry, Thomas Mendy |
| Donna Russell Consulting | Donna Russell |
| **Co-Investigator team** |  |
| Midlands State University, Zimbabwe | Prestige Tatenda Makanga, Liberty Makacha |
| King’s College London | Lucilla Poston, Rachel Tribe, Sophie Moore, Tatiana Salisbury |
| University of Oxford | Aris Papageorghiou, Alison Noble, Rachel Craik |
| London School of Hygiene and Tropical Medicine | Hannah Blencowe, Veronique Filippi, Joy Lawn, Matt Silver, Joseph Akuze and Ursula Gazeley |
| St George’s, University of London | Judith Cartwright, Guy Whitley, Sanjeev Krishna |
| University of British Columbia | Marianne Vidler, Jing (Larry) Li, Jeff Bone, Mai-Lei (Maggie) Woo Kinshella, Ash Sandhu |
| Imperial College London | Benjamin Barratt |
| University of Michigan, USA | Akbar Waljee |

## Supporting Information

**S1 Table: Method of gestational age assignment in the PRECISE cohort in sub-Saharan Africa [17].**

**\***Biometric measurements were crown-rump length, head circumference, and femur length. EDD, Estimated Date of Delivery; GA, gestational age; LMP, last menstrual period; SFH, symphyseal fundal height; US, ultrasound.

**S2 Table: Summary Statistics of Socio-demographic Variables and Other Risk Factors for Macrosomia from the PRECISE cohort in The Gambia, Kenya, and Mozambique.**

LBW, low birthweight; IQR, Interquartile range; DDS, dietary diversity score; Q1-Q4, quartiles 1-4. * Miscarriages (Gambia=12, Kenya=5, Mozambique=17) were excluded. †Number does not include the 34 missing birthweights from miscarriage pregnancies. **Exclusions related to birthweights are not mutually exclusive; for example, some multiple gestations are normal birthweight babies, others are LBW or macrosomia.

**S3 Table: Summary Statistics of Neonatal Outcome Variables for Macrosomia from the PRECISE (PREgnancy Care Integrating translational Science Everywhere) Pregnancy Cohort in The Gambia, Kenya and Mozambique.**

IQR, Interquartile range. Miscarriages and multiple gestations were excluded from this analysis.

**S4 Table: Stratification Analysis of Fetal Outcomes According to Macrosomia Severity (< 4500 g and ≥4500 g) from the PRECISE (PREgnancy Care Integrating translational Science Everywhere) Pregnancy Cohort in The Gambia, Kenya and Mozambique.**

OR, Odds Ratio; CI, confidence interval. * 3 missing data and ** each has 15 missing data; therefore, the percentages did not add up to 100%.

**S5 Table: Sub-country Analysis of Fetal Outcomes of Macrosomia from the PRECISE (PREgnancy Care Integrating translational Science Everywhere) Pregnancy Cohort in The Gambia, Kenya and Mozambique.**

Freq, frequency; OR, Odds Ratio; CI, confidence interval.

